# Predicting survival time for critically ill patients with heart failure using conformalized survival analysis

**DOI:** 10.1101/2024.09.07.24313245

**Authors:** Xiaomeng Wang, Zhimei Ren, Jiancheng Ye

**Author notes:** Corresponding author: Jiancheng Ye, PhD, Weill Cornell Medicine, Cornell University, New York, NY, USA.

## Abstract

Heart failure (HF) is a serious public health issue, particularly for critically ill patients in intensive care units (ICUs). Predicting survival outcomes of critically ill patients with calibrated uncertainty calibration is a difficult yet crucially important task for timely treatment. This study applies a novel approach, conformalized survival analysis (CSA), to predicting the survival time to critically ill HF patients. CSA quantifies the uncertainty of point prediction by accompanying each predicted value with a lower bound guaranteed to cover the true survival time. Utilizing the MIMIC-IV dataset, we demonstrate that CSA delivers calibrated uncertainty quantification for the predicted survival time, while the methods based on parametric models (e.g., Cox model or the Accelerated Failure Time model) fail to do so. By applying CSA to a large, real-world dataset, the study highlights its potential to improve decision-making in critical care, offering a more nuanced and accurate tool for prognostication in a setting where precise predictions and calibrated uncertainty quantification can significantly influence patient outcomes.

## 1 Introduction

Heart failure (HF) represents a significant and growing public health concern, with an estimated prevalence exceeding 64 million people worldwide [1]. As a chronic, progressive condition, HF is characterized by the heart’s inability to pump sufficient blood to meet the body’s needs, leading to a cascade of clinical manifestations that severely impair patients’ quality of life. Critically ill patients with HF represent a unique and particularly vulnerable subset of the broader HF population [2]. These patients are often admitted to the intensive care unit (ICU) due to acute decompensation or the presence of life-threatening complications such as cardiogenic shock, arrhythmias, or multi-organ failure [3]. The management of HF in this setting requires a nuanced understanding of the disease’s pathophysiology, as well as the ability to rapidly adjust treatment strategies in response to the patient’s evolving clinical status. Predicting outcomes in critically ill patients with HF is inherently difficult due to the dynamic nature of their condition [4]. The trajectory of HF in these patients can be influenced by a myriad of factors, including underlying comorbidities, the presence of acute precipitating events, the response to therapeutic interventions, and the patient’s overall functional status prior to ICU admission. The limitations of traditional prognostic models have spurred interest in more sophisticated approaches, particularly those that can accommodate the complexities and uncertainties inherent in HF. One such approach is survival analysis, a statistical method that deals with time-to-event data, where the event of interest could be death, hospitalization, or another adverse outcome. Traditional survival analysis methods, such as the Cox proportional hazards model and Kaplan-Meier curves, have been widely used to predict outcomes in HF patients [5]. However, these models have significant limitations when applied to critically ill populations. One major limitation is the assumption of proportional hazards, which may not hold true in the ICU setting where the patient’s risk of death can fluctuate rapidly based on their response to treatment and the development of new complications [5].

To address these challenges, this study utilizes a novel approach, conformalized survival analysis (CSA), which was first introduced in [6]. This method enhances traditional survival analysis by incorporating conformal prediction techniques, which allow for the generation of prediction intervals rather than single-point estimates. These prediction intervals provide a range of possible survival times with a specified level of confidence, offering a more nuanced and realistic assessment of the patient’s prognosis. Conformalized survival analysis is particularly well-suited to the ICU environment, where the ability to manage uncertainty is critical [6]. By providing clinicians with a range of likely outcomes, this model supports more informed decision-making, helping to guide the intensity of interventions and the allocation of resources based on the patient’s individualized risk profile [7]. Moreover, the use of prediction intervals helps to communicate the inherent uncertainty in survival predictions, which is particularly important in discussions with patients and their families about prognosis and treatment options.

This study aims to evaluate the effectiveness of conformalized survival analysis in predicting survival times for critically ill patients with HF. By applying this novel method to a large, real-world dataset of ICU patients, this study aims to demonstrate the advantages of conformalized prediction over traditional models in terms of accuracy, reliability, and clinical utility. The study also seeks to explore the potential implications of this approach for improving patient care in critical care settings, particularly in terms of personalized treatment planning and resource allocation.

## 2 Methods

### 2.1 Participants/Data

This study utilizes data from the MIMIC-IV (Medical Information Mart for Intensive Care IV) v2.2 dataset, a publicly available, large-scale, and de-identified dataset that contains detailed clinical data from patients admitted to ICUs at the Beth Israel Deaconess Medical Center in Boston, Massachusetts [8]. We included all patients who had at least one ICU admission (*n* = 43, 580). From this cohort, we filtered patients who had a diagnosis related to heart failure based on ICD codes. Subsequently, we selected only those patients who survived their ICU and hospital stay (whose date of death (dod) is not the same date as the last hospital admission discharge time), as censoring time is only available for these individuals. We excluded one patient due to incorrect data (date of death preceding the last ICU admission date). The final dataset consisted of *n* = 11, 712 patients. Survival time is defined with the last hospital discharge time as time zero (*T* = 0 days). The dataset contains follow up after hospital discharge for one calendar year, therefore, the censoring time for all patients is the same – 365 days. We used days as our unit because time of death is only available on the scale of days. For age, if a patient is over 89 years old, the age in the dataset is set to 91, regardless of the patient’s true age.

### 2.2 Exploratory Analysis

We conducted an exploratory analysis of predictor importance using Shapley values (referred to as SHAP values hereafter) [9]. The SHAP value measure the importance of a predictor by quantifying the contribution of it in a pre-specified model.

In the exploratory analysis, we used the binary indicator of whether patients died before the censoring time (*T <* 365 days) as the response variable and a set of features chosen clinically as the predictors. That is, we want to identify features that are most influential for whether a patient dies within one year of hospital discharge.

The full set of features chosen are: length of the latest ICU stay, demographic information (insurance, language spoken, gender, race, age, marital status), previous heart failure diagnosis (true if the patient’s heart failure diagnosis is from previous admissions, not the last ICU admission), comorbidities (hypertension, atrial fibrillation, Type-1 diabetes, Type-2 diabetes, Chronic Obstructive Pulmonary Disease (COPD), asthma, liver disease, chronic kidney disease, cancer, depression, anemia), and medication history (ACE inhibitors, beta-blockers, diuretics, anticoagulants).

### 2.3 Statistical Model

We employ the CSA framework introduced by [6]. To set the stage, let *T* represent the true survival time of a patient, *C* the censoring time, and *T̃* = min(*C, T*) the censored survival time. The observable data for patient *i* is denoted as the tuple (*X_i_, C_i_, T̃_i_*), where *X_i_* is a vector of predictors listed in Section 2.2.

The goal of CSA is to construct a lower bound for the true survival time *T* for a new patient based on *X*. Formally, we want to find *L̂*(*X*) such that

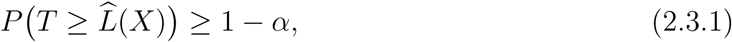

where *α* is a pre-specified significance level (e.g., 0.05).

In general, CSA addresses the censoring issue by fixing a threshold *c*_0_ and focusing on the subpopulation with *C ≥ c*_0_. In our specific application, the censoring time is a constant for all patients (*C ≡* 365). Therefore, we choose *c*_0_ = 365 and include all patients in our analysis.

We start by splitting the data into three folds: a training fold *Ƶ*_tr_, a calibration fold *Ƶ*_ca_, and a test fold *Ƶ*_test_, with a size 4000, 4000, and 3712 respectively. On *Ƶ*_tr_, we adopt the distribution boosting method [10] to fit a model for predicting the survival time. CSA then proceeds by computing a “conformity score” *V_i_* = *V* (*X_i_, Tı_i_*) for *i ∈ Ƶ*_ca_, where *V_i_* measures how the predicted survival time “conforms” to the true survival time. The final output lower bound is *L̂*(*x*) = inf*{y* : *V* (*x, y*) *≤ η*(*x*)*} ∧ c*_0_, where

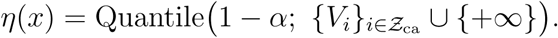

It has been proved that *L̂*(*x*) achieves the guarantee in (2.3.1). We refer the readers to [6] for the detailed theoretical results.

### 2.4 Comparison with other existing models

In Section 3.4, we compare our method with a list commonly used methods in survival analysis in their performance of uncertainty quantification. The benchmarks include the *Cox proportional hazard (Cox) model* [11], the *Accelerated failure time (AFT) model* [12, 13], two variants of censored quantile regression (CQR) by Powell [14] and Portnoy [15], and *censored quantile regression forest (RF)* [16]. In our comparison, Cox and AFT are implemented with the survival R-package [17]; the two variants of censored quantile regression are implemented with the quantreg R package [18], and censored RF with the code adapted from https://github.com/AlexanderYogurt/censored_ExtremelyRandomForest.

In our comparison, we evaluate the prediction sets provided by each method, checking empirically whether they achieve the target coverage level (a measure of validity) and their sizes (a measure of efficiency).

We note that both Cox and AFT are (semi-)parametric models, that assume the survival time depends on the predictors in specific ways. In these two methods, the prediction sets are derived based on the model assumptions, and their validity hinge on the correctly specifying the model and accurately estimating the parameters in the model. The two variants of censored quantile regression also makes parametric assumptions on how the quantiles depend on the predictors (e.g., linearly), and as a result, the corresponding prediction sets can be invalid when the assumptions are violated. Censored RF is a non-parametric method, and the validity of its prediction set is asymptotic and requires a set of non-parametric assumptions (e.g., Lipschitz conditions).

Although the above models have been popular choices for survival analysis, they all have implicit or explicit assumptions, on which the validity of the prediction sets hinges on. These assumptions are, in the first place, hard to verify. In addition, they are hardly satisfied in high-dimensional settings. In the case of model mis-specification (for example, when the we have second order effects), these models can produce large errors, as we will see in Section 3.4. Moreover, the validity of these methods are asymptotic, prone to violation when the sample size is small in practice.

Our method, on the other hand, is agnostic to model assumptions and finite-sample valid. That means, whether these model assumptions are satisfied or how many data points we have does not affect the validity of our prediction set. Therefore, our model is more robust to model mis-specification and small sample size.

### 2.5 Study Approval

This study exclusively used publicly available MIMIC-IV data.

## 3 Results

### 3.1 Patient Characteristics

We compared the characteristics of ICU patients including various demographic, clinical, and comorbidity characteristics between heart failure (HF) and non-heart failure (non-HF) patients. The results are presented in Table 1. We conducted two-sided t-tests for all continuous variables and chi-square tests for all categorical variables to assess the differences in these characteristics between the two groups. The p-values for these tests are listed in the last column. In all demographic features and all comorbidities except for liver disease and chronic kidney disease, the two group had significant differences (*p <* 0.05).

**Table 1:** Comparison of characteristics between heart failure and non-heart failure groups

| Variables | Total | Heart failure | Non-heart failure | P-value |
| --- | --- | --- | --- | --- |
| **Sex** (Male) | 56.1% | 54.9% | 56.5% | 0.00222 |
| **Language** (English) | 90.3% | 89.3% | 90.7% | <0.0001 |
| **Insurance** |  |  |  |  |
| Medicare | 42.0% | 59.1% | 35.7% | <0.0001 |
| Medicaid | 15.8% | 36.7% | 8.1% |  |
| Other | 42.2% | 4.2% | 56.1% |  |
| **Marital Status** |  |  |  |  |
| Married | 45.9% | 44.7% | 46.4% | <0.0001 |
| Single | 27.7% | 22.2% | 29.7% |  |
| Widowed | 11.8% | 20.1% | 8.7% |  |
| Divorced | 7.2% | 7.6% | 7.1% |  |
| **Age** (mean, sd) | (64, 17.3) | (72, 13.7) | (61, 17.5) | <0.0001 |
| **Race** |  |  |  |  |
| White | 68.7% | 71.8% | 67.5% | <0.0001 |
| Black | 9.2% | 10.6% | 8.7% |  |
| Hispanic | 3.7% | 3.2% | 3.8% |  |
| Asian | 2.9% | 2.1% | 3.2% |  |
| Other<sup>1</sup> | 15.6% | 12.4% | 16.8% |  |
| **Comorbidities** |  |  |  |  |
| Hypertension | 25.1% | 20.2% | 26.9% | <0.0001 |
| Atrial fibrillation | 10.8% | 21.6% | 6.8% | <0.0001 |
| Type 1 diabetes | 1.2% | 1.4% | 1.2% | 0.02852 |
| Type 2 diabetes | 9.5% | 18.4% | 6.2% | <0.0001 |
| COPD | 7.4% | 14.3% | 4.8% | <0.0001 |
| Asthma | 5.6% | 7.1% | 5.0% | <0.0001 |
| Liver disease | 0.2% | 0.2% | 0.2% | 0.41222 |
| Chronic kidney disease | 0.3% | 0.3% | 0.3% | 0.7414 |
| Cancer | 2.0% | 2.6% | 1.8% | <0.0001 |
| Depression | 13.3% | 16.9% | 11.9% | <0.0001 |
| Anemia | 12.5% | 16.6% | 11.0% | <0.0001 |
| **Death**, N (%) | 21.0% | 34.8% | 15.9% | <0.0001 |

<sup>1</sup> Other includes all other categories that are not White, Black, Hispanic, and Asian. For example, this includes all mixed race, Pacific Islander, Native American, and unknowns.

**Table 2:** Comparison of quantiles of predicted lower bound length for various methods

| Quantile | Cox | AFT | Powell | Portnoy | RF | CSA |
| --- | --- | --- | --- | --- | --- | --- |
| Q1 | 19.79 | 15.63 | 22.20 | 22.50 | 19.81 | 17.98 |
| Median | 27.17 | 27.91 | 35.23 | 35.88 | 27.02 | 27.94 |
| Q3 | 40.98 | 53.89 | 50.58 | 51.87 | 38.89 | 50.93 |

The HF group tends to be older (mean age 72 years vs. 61 years) and has a higher prevalence of comorbidities such as atrial fibrillation (21.6% vs. 6.8%), Type 2 diabetes (18.4% vs. 6.2%), and COPD (14.3% vs. 4.8%). Insurance status also significantly differs, with a larger proportion of HF patients covered by Medicare (59.1% vs. 35.7%) and Medicaid (36.7% vs. 8.1%). Marital status shows that HF patients are more likely to be widowed (20.1% vs. 8.7%), which is likely correlated with the older age profile. Racial differences are relatively minimal, but the non-HF group has a slightly higher percentage of patients who speak English as their primary language (90.7% vs. 89.3%). Importantly, the mortality rate is significantly higher in the HF group (34.8% vs. 15.9%). Almost all comparisons yield statistically significant differences (before multiple comparison correction, p *<* 0.05), highlighting the distinct characteristics and higher risk profile of the HF group compared to non-HF patients.

### 3.2 Univariate predictors of survival

The SHAP summary plot in Figure 1 provides a clear visualization of how various factors influence the likelihood of a patient dying within a year after hospital discharge. Age emerges as the most significant predictor, with older patients (indicated by red dots) having a general higher risk of death. The length of ICU stay is another important factor, where longer stays are associated with an increased risk. In contrast, the use of ACE inhibitors appears to reduce the risk of death, particularly for patients with higher medication usage (indicated by blue dots). Heart failure patients with hypertension may live longer than those with heart failure alone; patients with hypertension may be diagnosed and treated earlier and receive medical intervention and closer monitoring, potentially leading to better outcomes. Insurance type also plays a role; patients with Medicare insurance generally show a higher probability of death.

**Figure 1:**
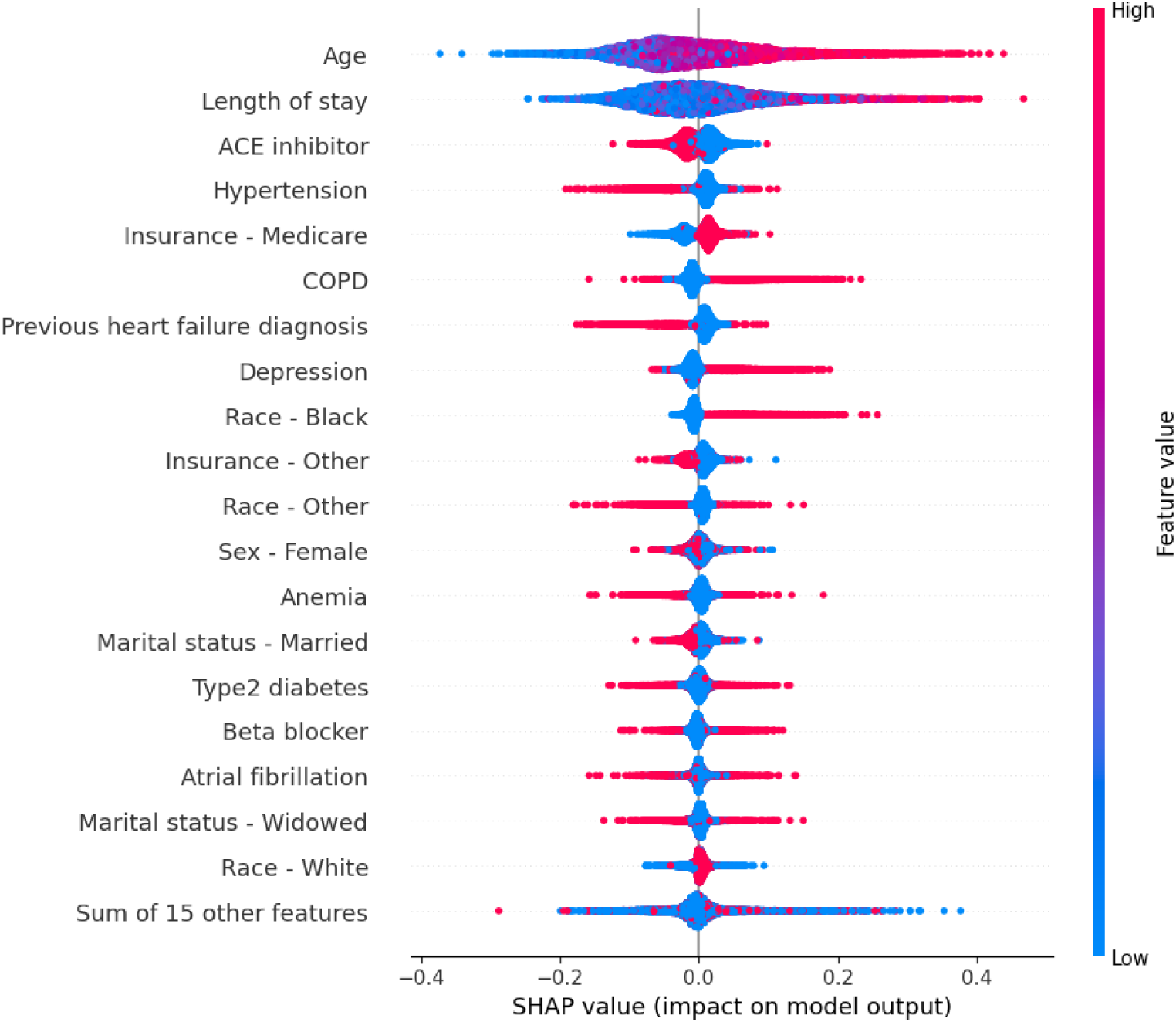
Shapley value for all predictors

### 3.3 Predicted lower bound of survival time

We randomly split the sample into two equal groups (*n*_1_ = *n*_2_ = 5, 856). For each group, we used the other group as training data to fit a model and then used this model to predict the survival time lower bound at *α* = 0.1. The results are presented in Figure 2, where the predicted lower bound is compared against sex, race, and age, respectively.

**Figure 2:**
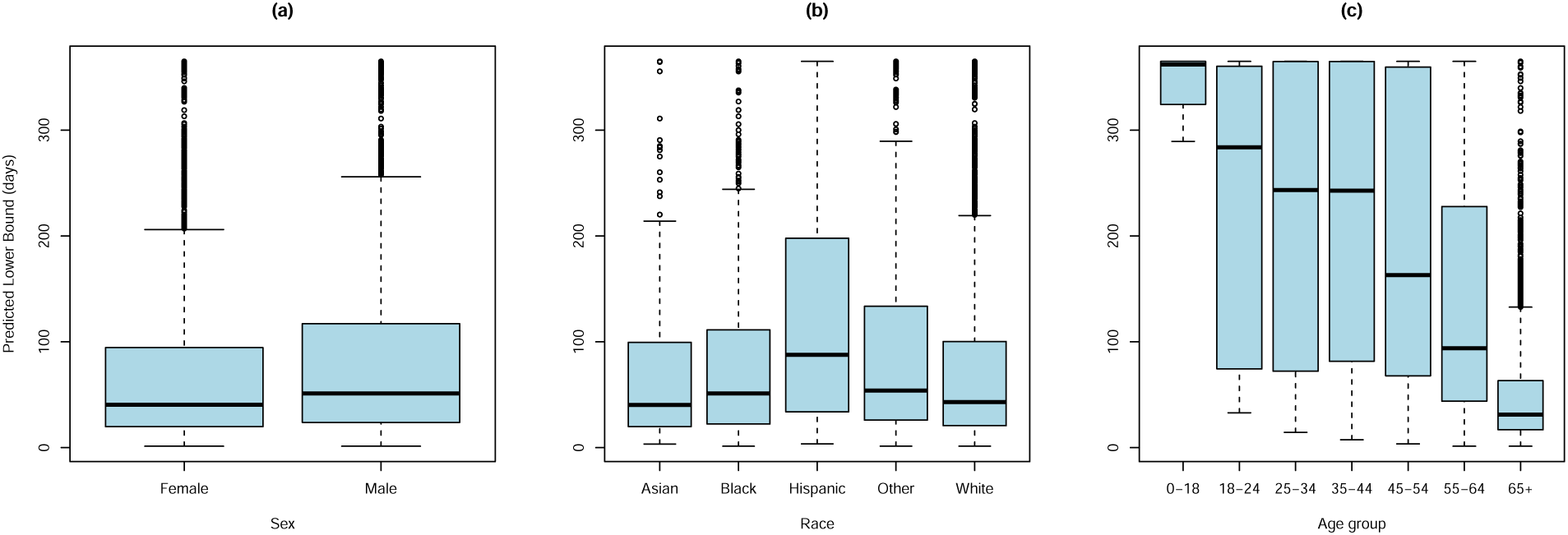
Predicted survival lower bound by: (a) sex; (b) race; (c) age group.

These figures are intended to visualize the predicted outcomes rather than to make formal statistical comparisons, as other predictors and potential confounders are present. However, given the sample size, it is reasonable to assume that other predictors are relatively balanced within each subgroup. Therefore, this visual comparison can still provide useful insights.

In Figure 2(a), we can see that the predicted lower bound is slightly higher for males than females. The lower 25% quantile are about the same for both genders but the 75% quantile is larger for male, suggesting that there’s more more males that have relatively high predicted lower bound.

In Figure 2(b), Hispanic showed the highest average survival lower bound. The other four categories have roughly the same mean, with Black and Other being slightly higher. This could be due to the difference in sample size. Table 1 shows that the dataset contains predominately white patients. Since the sample sizes for other categories are much smaller, the variance will be larger for these categories.

In the comparison for different age groups in Figure 2(c), there is a clear trend of survival time lower bound decreasing as age increases. For the 0-18 age group, all patients had a predicted lower bound of about a year. Then the median of the lower bound decreased to just above 250 days for the 18-24 age group, and stayed above 200 days for 25-34 and 35-44 age groups. After that, there is a consistent significant decrease for every age group. For ages 45-54, the median decreased to below 200 days, and for ages 55-64, the median decreased to about 100 days. For the age group 65+, the median is less than 50 days.

### 3.4 Empirical coverage

We compared the empirical coverage of our method to the traditionally used methods introduced in Section 2.4. We used *n* = 8000 data as the training set and *n* = 3712 as testing set.

We compared empirical coverage in both low-dimensional and high-dimensional settings. In the low-dimensional setting, we used *p* = 23 predictors as described in Section 2.3. In the high-dimensional setting, we used the same 23 predictors along with 100 additional binary predictors for whether the patients have been prescribed 100 different kind of medications to test if the performance of our method is robust in high-dimensional settings. For both cases, we tested three different *α* levels: 0.1, 0.05 and 0.01. For each method, we randomly split the training-testing data and repeat the experiment 250 times to get an empirical distribution of the coverage.

The empirical coverage of all methods in the low-dimensional case is shown in Figure 3. For each *α*, we would like to see the empirical coverage to be as close to 1 *− α* as possible. A low coverage (*<* 1 *− α*) would mean that the method fails to provide valid coverage guarantee, while a high coverage (*>* 1 *− α*) would mean that although the method can cover the true value most of the times, it is being over-conservative. Both cases are undesirable in application.

**Figure 3:**
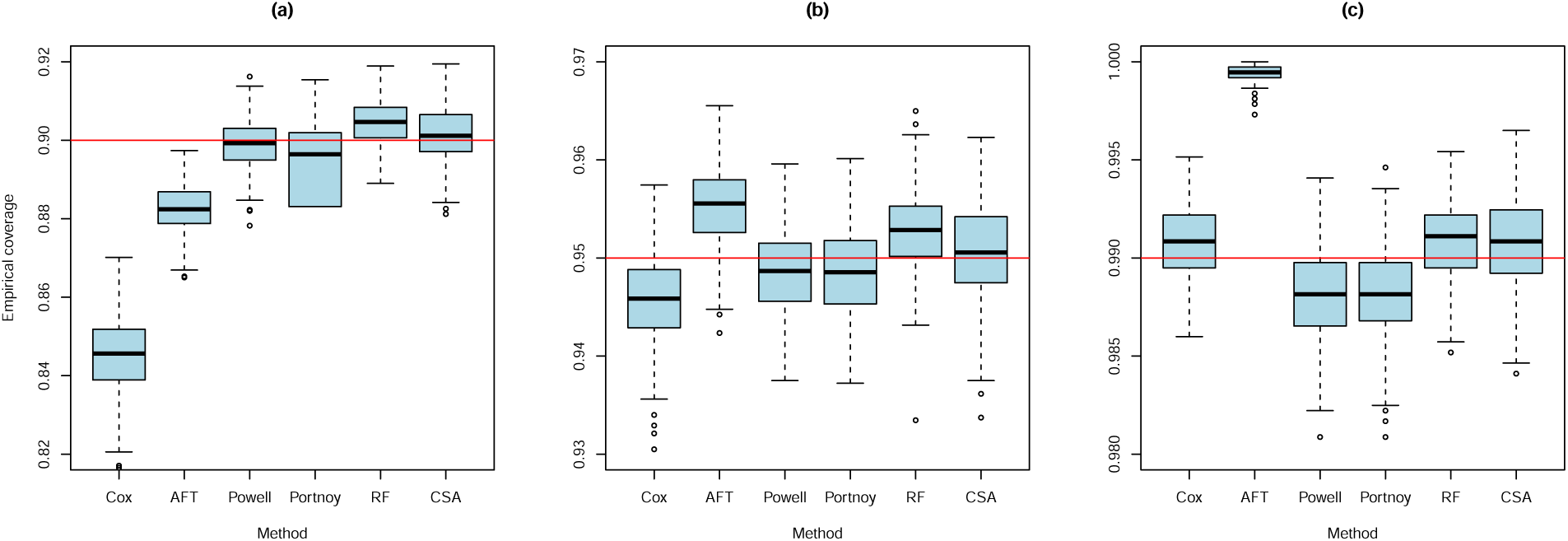
Empirical coverage in low-dimensional setting for: (a) *α* = 0.1; (b) *α* = 0.05; (c) *α* = 0.01. The methods used are Cox proportional hazard model (Cox), Accelerated failure time (AFT), Censored quantile regression (Powell and Portnoy), Censored quantile regression forest (RF), and Conformalized survival analysis (CSA). The red horizontal line is the target coverage.

We can see that our method consistently produces the desired coverage without being over-conservative. Cox and AFT under-cover for large *α* and over-cover for small *α*. The two censored quantile regression variants consistently under-cover while RF consistently over-cover.

The empirical coverage in the high-dimensional case is shown in Appendix A.1. Again, our method is the only method providing consistent target coverage across different *α* levels. Among the rest of the methods, we can see that our method is still robust in the high-dimensional setting, producing valid and non-conservative predictions.

In additional to the marginal coverage, we also examined the conditional coverage by sex, race, and age group in the low-dimensional setting with *α* = 0.05 as an example. The results are shown in Appendix A.1. Although our method does not provide conditional coverage guarantee, we can see through this set of figures that our method has relatively consistent coverage across different sub-population. However, the most popular Cox method has very different conditional coverage in different sub-population, which is sub-optimal in most cases.

### 3.5 A survival lower bound calculator

We provide a survival lower bound calculator that is publicly accessible online at https://username434.shinyapps.io/Heart_failure_conformalized_survival_analysis/. A screenshot of the user interface is shown in Figure 4. Users can use this interface by: (1) inputting the demographic information and medical history of any patient; (2) inputting a desired *α ∈* [0, 1] level, and (3) click the **calculate** button to calculate a *α*-level survival lower bound prediction for the specified *α* value using our pre-trained model. We also provide a plot of survival lower bound prediction for *α ∈* [0.02, 0.5] to help visualize the prediction, Figure 5 is shown as an example. In the example below, this patient’s survival lower bound is about 100 days for *α* = 0.1 and about 250 days for *α* = 0.2. This suggests that we can predict with 90% confidence that this patient can survive longer than 100 days and with 80% confidence this patient can survive longer than 250 days. However, we want to make the clarification that our method has coverage guarantee for only a specific pre-fixed *α*. It does not have a marginal guarantee for all *α* levels jointly.

**Figure 4:**
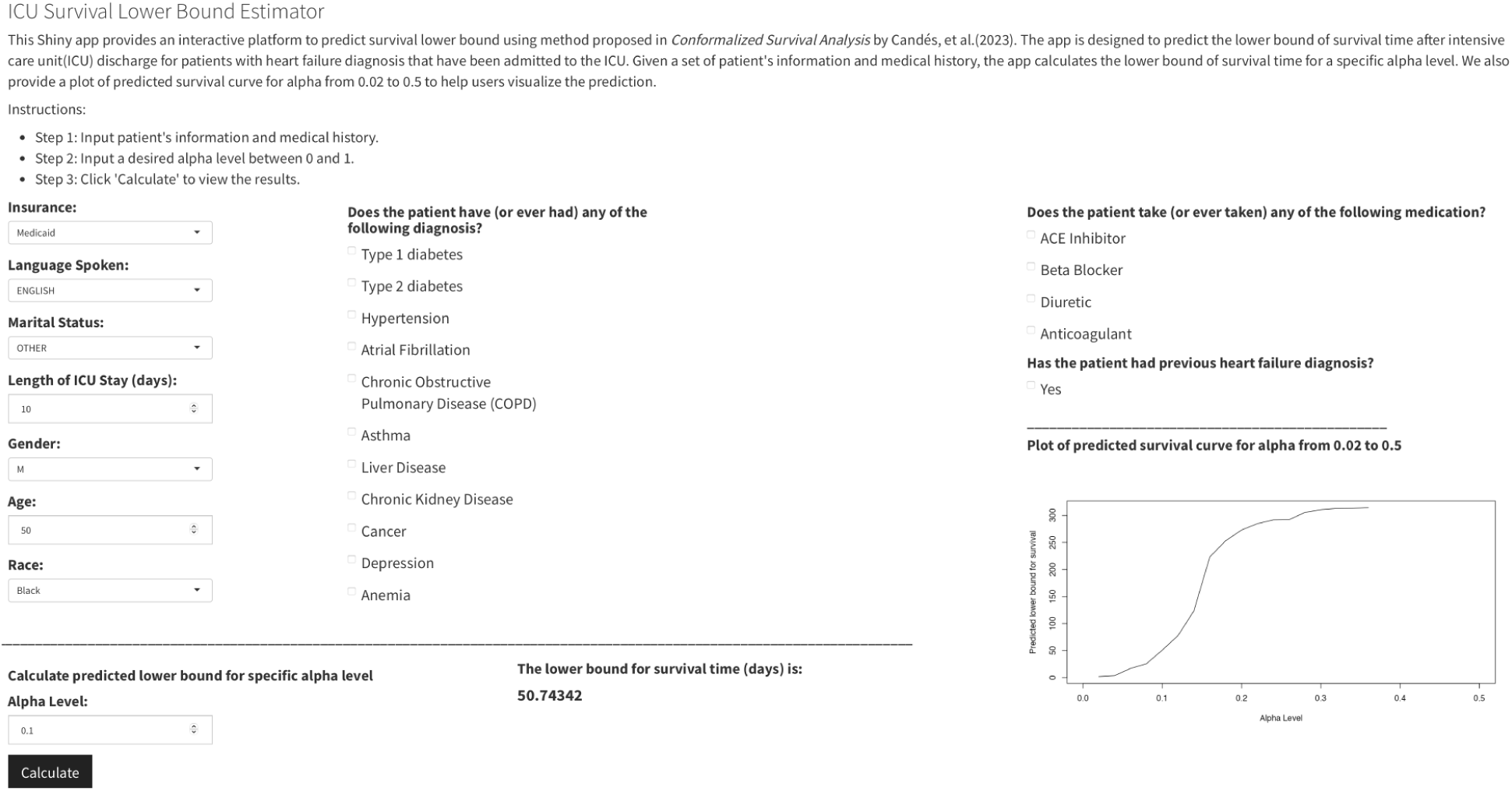
Online survival lower bound calculator dashboard

**Figure 5:**
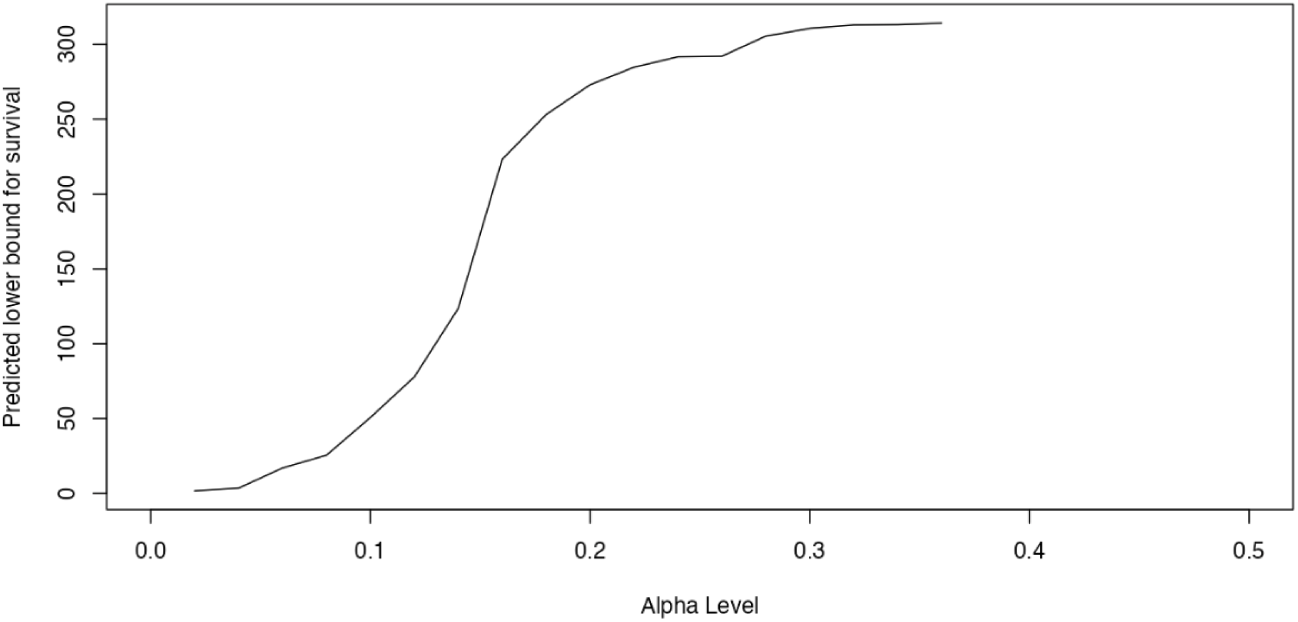
Online survival lower bound calculator example plot

## 4 Discussion

This study explored the application of conformalized survival analysis (CSA) to predict survival times for critically ill patients with heart failure. By leveraging this novel approach, we aimed to provide reliable and interpretable prediction intervals that can support decision-making processes in the high-stakes environment of critical care. Our findings demonstrate that CSA offers several advantages over traditional survival analysis methods, including improved accuracy and the ability to quantify uncertainty.

The application of CSA to predict survival times in critically ill patients with HF represents a significant advancement in the field of critical care prognostication. CSA outperformed traditional models, such as the Cox proportional hazards model, particularly in its ability to account for the variability in patient outcomes. The inclusion of prediction intervals adds a crucial layer of information, allowing clinicians to assess the confidence of each prediction and adjust their clinical decisions accordingly [19]. The results presented in this study underscore the method’s ability to generate accurate and reliable predictions, even in the highly complex and variable context of critically ill patients. Specifically, the model demonstrated high levels of accuracy in predicting survival times across different time intervals, with prediction intervals providing clinicians with a clear range of likely outcomes. This study’s findings are particularly relevant for critically ill patients, as the model’s predictions can inform critical decisions about the intensity and direction of care.

Critically ill patients with HF often present a significant challenge due to the complex interplay of factors that influence their prognosis. These patients frequently exhibit rapid changes in their clinical condition, making it difficult to predict outcomes using traditional models that rely on static assumptions. The study’s findings are especially relevant in this context, as CSA offers a dynamic approach to prognostication that better reflects the realities of critical care [20]. This study showed that the model’s prediction intervals were particularly useful in capturing the uncertainty inherent in the care of critically ill patients. For instance, in cases where patients exhibited fluctuating clinical parameters, the model provided broader prediction intervals, reflecting the increased uncertainty. This feature is critical in the ICU setting, where decisions about escalating or de-escalating care must often be made with incomplete information. By providing a range of potential outcomes, CSA allows clinicians to make more informed decisions that consider both the best-case and worst-case scenarios.

Traditional survival models, such as the Cox proportional hazards model, have been used for predicting outcomes in HF patients. However, these models have several limitations when applied to critically ill populations [21]. In contrast, CSA provided more nuanced predictions that reflected the dynamic nature of critical illness. Another key finding was the method’s ability to generate more accurate predictions for patients who exhibited rapid changes in their clinical status. Traditional models tend to struggle in such scenarios, as they rely on assumptions that may not hold true in a rapidly changing environment. CSA, by contrast, was able to adapt to these changes, providing predictions that better reflected the patient’s current condition. This adaptability is particularly important in the ICU, where timely and accurate predictions can significantly impact patient outcomes.

The conformalized prediction intervals were narrower and more precise, allowing for better risk stratification and individualized patient care [20]. The use of machine learning techniques in combination with conformal prediction enabled the model to capture complex interactions among variables, which is essential for the heterogeneous and dynamic nature of critically ill patients with HF [6]. In addition, CSA has strength in the robustness and interpretability. This method provides valid prediction intervals that are robust to model mis-specification, offering clinicians a quantifiable measure of confidence in the predictions. We have shown that CSA can consistently provide valid prediction despite model size for any fixed *α* level, robust to modeling assumptions. This is particularly important in the ICU setting, where decisions often need to be made quickly with confidence using incomplete information. The interpretability of CSA also enhances its clinical utility, as it allows healthcare providers to understand the factors influencing the predictions and to make informed decisions based on the predicted outcomes [22].

The clinical implications of these findings are profound. In the ICU, where decisions must often be made rapidly and with limited information, having access to reliable and individualized survival predictions can greatly enhance the quality of care. For critically ill HF patients, these clinical decisions could mean the difference between life and death, as well as the difference between receiving care that is appropriately aggressive versus care that is overly burdensome [23]. The ability to generate prediction intervals is particularly valuable in this context, as it allows clinicians to assess the likelihood of different outcomes and adjust their treatment plans accordingly. For example, a patient with a narrow prediction interval indicating a high likelihood of survival may be a candidate for aggressive treatment, while a patient with a broader interval indicating greater uncertainty may benefit from a more conservative approach [24]. This level of precision in prognostication can help to optimize the use of ICU resources, ensuring that patients receive the level of care that is most appropriate for their situation. Moreover, the findings have important implications for communication with patients and their families. In the ICU, discussions about prognosis are often fraught with uncertainty, which can lead to misunderstandings and difficult decisions. CSA can help to clarify these discussions by providing a range of possible outcomes and the associated confidence levels. This information may facilitate more informed and collaborative decision-making, ensuring that treatment plans align with the patient’s goals and preferences [25].

### Limitations

While our study demonstrates the potential benefits of CSA in predicting heart failure survival times, there are several limitations that need to be addressed. Firstly, the study was conducted using data from a single dataset (MIMIC-IV v2.2), and the results may not be generalizable to other populations or settings. Future studies should validate the findings using data from different sources and diverse patient populations. Secondly, the study focused on predicting survival times up to one year after ICU admission, and further research is needed to explore the applicability of CSA for longer-term predictions. Lastly, while CSA provides valid prediction intervals, the method relies on the availability of high-quality data. Ensuring the accuracy and completeness of the input data is essential for obtaining reliable predictions.

## 5 Conclusion

Our study highlights the potential of conformalized survival analysis as a powerful tool for predicting survival times in heart failure patients admitted to the ICU. The method offers several advantages over traditional survival analysis techniques, including improved predictive accuracy, robustness, and interpretability. By providing reliable and quantifiable prediction intervals, CSA can support clinical decision-making and enhance patient care in the critical care setting. Future research should focus on validating the findings in diverse populations and exploring the applicability of CSA for longer-term predictions. The integration of advanced machine learning techniques with conformal prediction represents a promising direction for improving prognostic models and optimizing patient outcomes in the ICU.

## Contribution Statement

XW and ZR contributed to data analyses and the writing of the manuscript. JY designed the study, contributed to the data analyses, and the writing of the manuscript. All authors read and approved the final version of the manuscript.

## Data Availability Statement

All data are available at Medical Information Mart for Intensive Care: https://mimic.physionet.org/. The relevant code and analyses are available at: https://github.com/Xiaomengwang99/Heart_Failure_CSA.

## Data Availability

All data are available at Medical Information Mart for Intensive Care: https://mimic.physionet.org/

## A Appendix

### A.1 Additional Experiment Results

Here we provide additional experiment results in the high-dimensional setting. The coverage for all methods compared is shown in Figure 6. Due to the scale of the figure, we also provide a zoomed-in version without the worst performing Cox and Portnoy methods. This version is shown in Figure 7.

**Figure 6:**
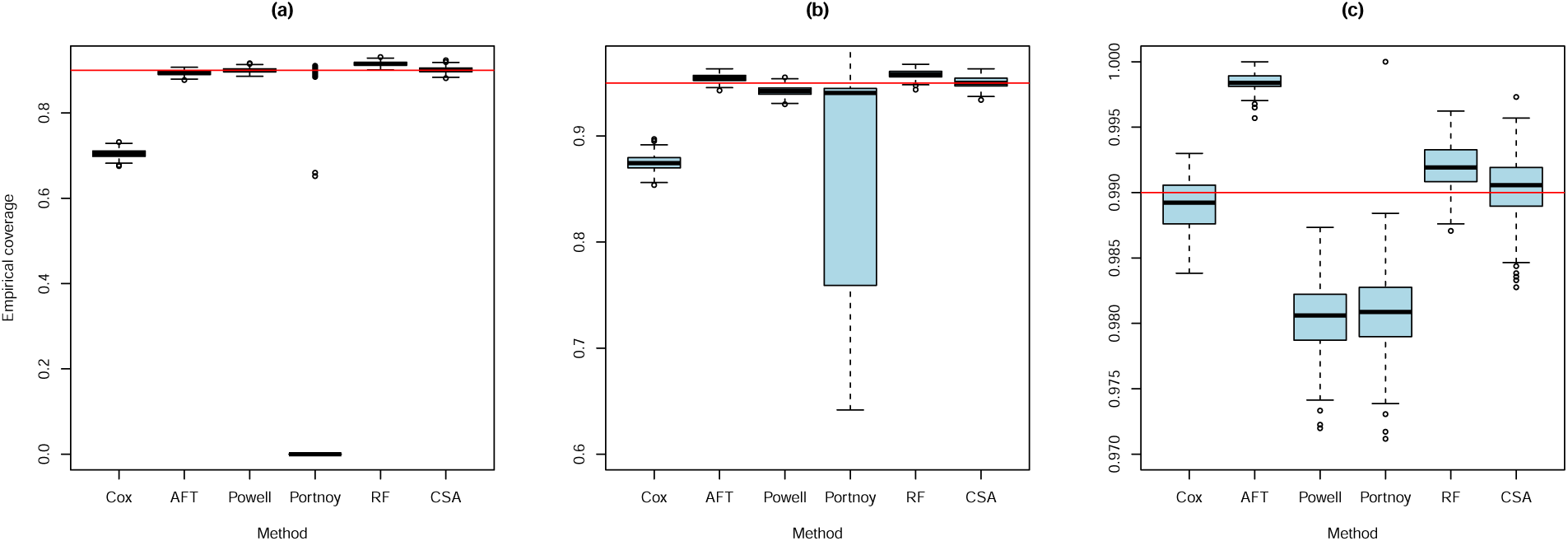
Empirical coverage in high-dimensional setting for: (a) *α* = 0.1; (b) *α* = 0.05; (c) *α* = 0.01. The methods used are Cox proportional hazard model (Cox), Accelerated failure time (AFT), Censored quantile regression (Powell and Portnoy), Censored quantile regression forest (RF), and Conformalized survival analysis (CSA). The red horizontal line is the target coverage.

**Figure 7:**
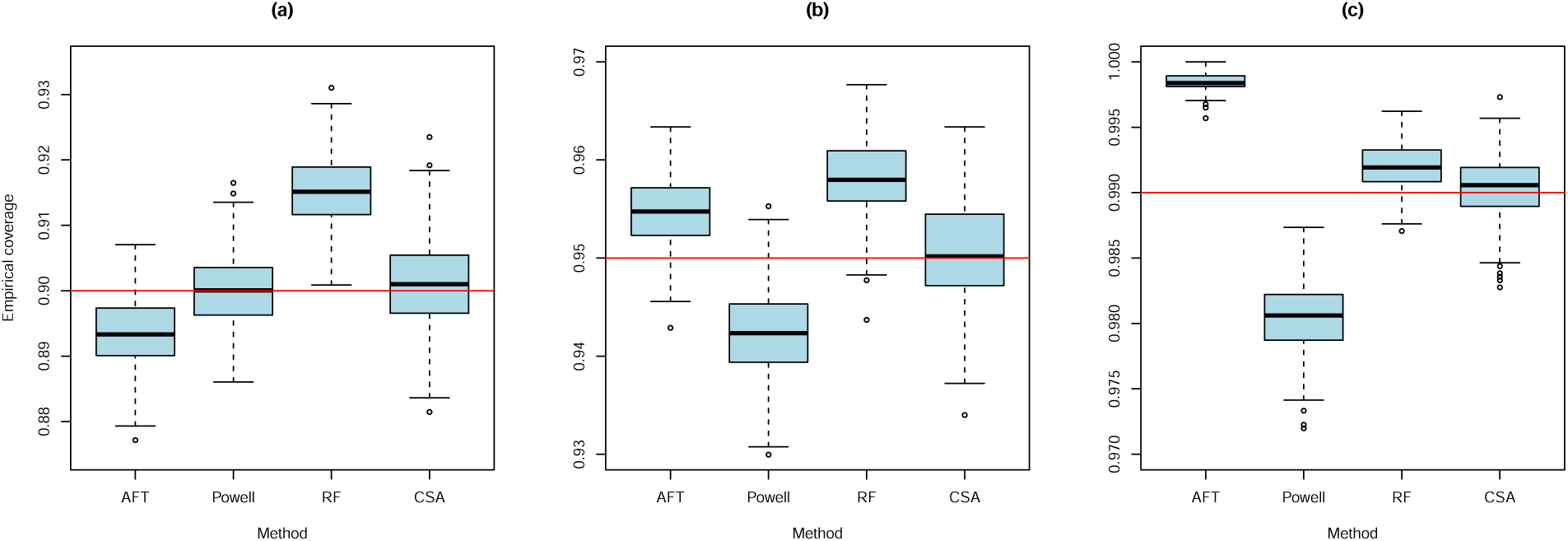
Empirical coverage in high-dimensional setting for: (a) *α* = 0.1; (b) *α* = 0.05; (c) *α* = 0.01. The methods used are Accelerated failure time (AFT), Censored quantile regression (Powell), Censored quantile regression forest (RF), and Conformalized survival analysis (CSA). The red horizontal line is the target coverage.

In Figure 8, we present the conditional coverage by sex, race, and age group produced by various methods.

**Figure 8:**
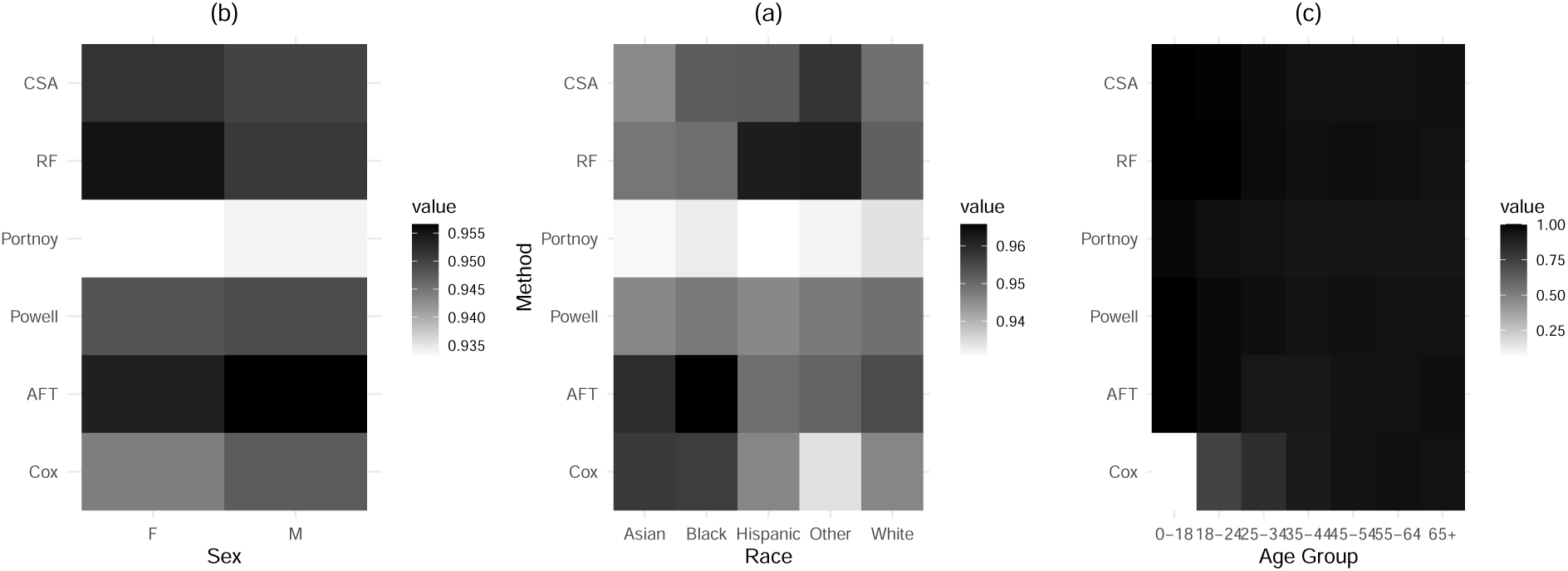
Conditional coverage by: (a) sex; (b) race; (c) age group. The methods used are Accelerated failure time (AFT), Censored quantile regression (Powell), Censored quantile regression forest (RF), and Conformalized survival analysis (CSA).

We also compared the he length of the predicted survival lower bound by different methods. The summary statistics are shown in Figure 2.

## References

[1] Gianluigi Savarese, et al. “Global burden of heart failure: a comprehensive and updated review of epidemiology”. In: Cardiovascular research 118.17 (2022), pp. 3272–3287.

[2] Owais Dar and Martin R Cowie. “Acute heart failure in the intensive care unit: epidemiology”. In: Critical Care Medicine 36.1 (2008), S3–S8.

[3] Khalid Bin Saleh et al. “Clinical characteristics and outcomes of patients with heart failure admitted to the intensive care unit with coronavirus disease 2019 (COVID-19): a multicenter cohort study”. In: American Heart Journal Plus: Cardiology Research and Practice 7 (2021), p. 100033.

[4] Shengxian Peng et al. “Interpretable machine learning for 28-day all-cause in-hospital mortality prediction in critically ill patients with heart failure combined with hypertension: A retrospective cohort study based on medical information mart for intensive care database-IV and eICU databases”. In: Frontiers in cardiovascular medicine 9 (2022), p. 994359.

[5] Asif Newaz, Nadim Ahmed, and Farhan Shahriyar Haq. “Survival prediction of heart failure patients using machine learning techniques”. In: Informatics in Medicine Unlocked 26 (2021), p. 100772.

[6] Emmanuel Candès, Lihua Lei, and Zhimei Ren. “Conformalized survival analysis”. In: Journal of the Royal Statistical Society Series B: Statistical Methodology 85.1 (2023), pp. 24–45.

[7] Jiancheng Ye, et al. “Predicting mortality in critically ill patients with diabetes using machine learning and clinical notes”. In: BMC medical informatics and decision making 20 (2020), pp. 1–7.

[8] A Johnson et al. MIMIC-IV (version 1.0). 2020.

[9] Scott M Lundberg and Su-In Lee. “A unified approach to interpreting model predictions”. In: Advances in neural information processing systems 30 (2017).

[10] Jerome H Friedman. “Contrast trees and distribution boosting”. In: Proceedings of the National Academy of Sciences 117.35 (2020), pp. 21175–21184.

[11] D. R. Cox. “Regression Models and Life-Tables (with discussion)”. In: Journal of the Royal Statistical Society: Series B (Methodological) 34.2 (1972), pp. 187–220.

[12] John D. Kalbfleisch and Ross L. Prentice. The Statistical Analysis of Failure Time Data. 2nd ed. John Wiley & Sons, 2002.

[13] L. J. Wei. “The Accelerated Failure Time Model: A Useful Alternative to the Cox Regression Model in Survival Analysis”. In: Statistics in Medicine 11.14-15 (1992), pp. 1871–1879.

[14] James L Powell. “Censored regression quantiles”. In: Journal of econometrics 32.1 (1986), pp. 143–155.

[15] Stephen Portnoy. “Censored regression quantiles”. In: Journal of the American Statistical Association 98.464 (2003), pp. 1001–1012.

[16] Alexander Hanbo Li and Jelena Bradic. “Censored quantile regression forest”. In: International Conference on Artificial Intelligence and Statistics. PMLR. 2020, pp. 2109–2119.

[17] Terry M Therneau. *A Package for Survival Analysis in R*. R package version 3.7-0. 2024. url: https://CRAN.R-project.org/package=survival.

[18] Roger Koenker, et al. “Package ‘quantreg’”. In: Reference manual available at R-CRAN: https://cran.rproject.org/web/packages/quantreg/quantreg.pdf (2018).

[19] Hans-Christian Thorsen-Meyer, et al. “Discrete-time survival analysis in the critically ill: a deep learning approach using heterogeneous data”. In: NPJ digital medicine 5.1 (2022), p. 142.

[20] Shubha Sankar Banerjee. “Conformalized Survival Analysis: A Review”. In: (2023).

[21] Eric D Adler et al. “Improving risk prediction in heart failure using machine learning”. In: European journal of heart failure 22.1 (2020), pp. 139–147.

[22] Yu Gui, et al. “Conformalized survival analysis with adaptive cut-offs”. In: Biometrika 111.2 (2024), pp. 459–477.

[23] Jiancheng Ye and L Nelson Sanchez-Pinto. “Three data-driven phenotypes of multiple organ dysfunction syndrome preserved from early childhood to middle adulthood”. In: AMIA Annual Symposium Proceedings. Vol. 2020. American Medical Informatics Association. 2020, p. 1345.

[24] Jiancheng Ye. “Patient safety of perioperative medication through the lens of digital health and artificial intelligence”. In: JMIR Perioperative Medicine 6 (2023), e34453.

[25] J Randall Curtis, et al. “Facilitating communication for critically ill patients and their family members: study protocol for two randomized trials implemented in the US and France”. In: Contemporary clinical trials 107 (2021), p. 106465.

